# Development and Validation of World Health Organization Assessment Schedule 2.0 for persons with disabilities having community rehabilitation services in Hong Kong

**DOI:** 10.1101/2025.06.01.25328755

**Authors:** Tiffany Ching Man Choi, Yin Kin Lee, Ho Lim Lee, Pandora Chao Ming Wang, Herman Mun Cheung Lau, Arran Siu Lun Leung, Eric Lu Shek Chan

## Abstract

**Background:** World Health Organization Disability Assessment Schedule 2.0 (WHODAS 2.0) was developed in 2010 to cover core components and constructs in ICF and assess disability. Simplified Chinese version and modified version by Taiwanese experts are not applicable for Hong Kong population. Therefore, there is a need for developing Hong Kong Chinese version of WHODAS 2.0 (WHODAS2.0 TC-HK). The aim of this study is to develop the WHODAS2.0 TC-HK and to evaluate its validity and reliability for persons with disabilities having community services.

**Methods:** WHODAS 2.0 was translated from the original English version to Hong Kong Chinese. 177 subjects with disabilities were recruited from the 4 community rehabilitation units subsidized by the government and held at a local non-governmental organization (NGO) to complete WHODAS2.0 TC-HK, Chinese HK version of 36-Item Short Form Health Survey (SF-36 TC-HK) and Hong Kong version of World Health Organization Quality of Life Abbreviated (WHOQOL-BREF TC-HK). The average age was 55.2 (SD 15.1) years, and 53.1% were women. Test-retest reliability, Internal consistency, Convergent validity, Concurrent validity and Construct validity were examined.

**Results:** Test-retest reliability and internal consistency of WHODAS2.0 TC-HK were moderate to good and good, respectively. In convergent and concurrent validity, WHODAS2.0 TC-HK domains were negatively and moderately correlated to SF-36 TC-HK sub-scores and WHOQOL-BREF TC-HK sub-scores. Confirmatory factor analysis (CFA) did not show satisfactory fit indices and the factor loadings per item were fair.

**Conclusions:** WHODAS2.0 TC-HK is a valid and reliable instrument in accessing disability based on ICF model for clients at community rehabilitation services in Hong Kong.

## Introduction

According to the International Classification of Functioning, Disability and Health (ICF), functioning and disability are a result of the interaction of an individual’s health condition and contextual factors (environmental and personal factors) [1]. The ICF classification of disability has shifted from the previous medically focused definition to a biopsychosocial approach [2]. This complex, dynamic, multidimensional, and contested nature of the concept of disability increases the challenges of disability measurement [3]. Whilst the ICF framework appreciates the interaction between different factors that affect disability and functioning, its complexity in linking the standard instrument of disability measurement to the ICF model conceptually and operationally makes direct implementation in clinical practice challenging [4]. A variety of ICF-based measurement tools were therefore derived, attempting to develop a standardised procedure to collect disability and functioning data. The World Health Organization Disability Assessment Schedule 2.0 (WHODAS 2.0) was invented in 2010 [5,6]. It is a 36-item self-administered tool designed to assess functioning in six life domains. The WHODAS 2.0 acted as a generalised and cross-cultural method to measure individuals’ limitations on different activities and social participation in a way that is independent of medical diagnosis [7]. It has been used in cross-country research for general populations and those with physical, mental, and neurological conditions [5,8,9].

The 36-item WHODAS 2.0 has been validated with adequate reliability in various groups, including arthritis, stroke, chronic psychotic disorder, depression, and back pain [10–13]. The WHODAS 2.0 supersedes WHODAS II and shows many advantages for example used across all diseases including mental, neurological and addictive disorders; short, simple and easy to administer; applicable in both clinical and general population settings; producing standardized disability levels and profiles; applicable across cultures, in all adult populations and directly linked at the level of the concepts to ICF [6].

WHODAS 2.0 has been translated into around 50 languages and dialects and is used in almost 30 areas of research in around 100 countries [6]. Experts in Taiwan once modified and translated WHODAS 2.0 to an instrument called Functioning Disability Evaluation Scale (FUNDES), which was in Traditional Chinese with an excellent internal consistency (Cronbach’s alpha: 0.9); however, it was not applicable in Hong Kong communities as that version included two new domains which were not shown in the English original version [14].

A simplified Chinese version of WHODAS 2.0 has also been developed. However, this version may not be completely applicable in Hong Kong since Cantonese (Hong Kong Chinese, in the form of Traditional Chinese) is the most common form/dialect of spoken Chinese used by the Hong Kong population, and there were cultural differences in language semantics, idiomatic expressions and concepts [15].

In Hong Kong, the government highly recommended the application of ICF in clinical assessment, collecting statistics, formulating rehabilitation plans, and prioritising social services as mentioned in the Persons with Disabilities and Rehabilitation Programme Plan in 2020[16]. Yet, there is no generic assessment tool revealing the disability status for local persons with disabilities in community services. Many assessment instruments mainly focus on functional activities such as walking, eating, dressing, and grooming instead of social participation and activities [8]. Also, service users cannot participate and express their concerns in the rehabilitation plan [17]. The adaptation of the WHO standardized measurement tool with the ICF framework in Hong Kong, WHODAS2.0 TC-HK, seems to be a more feasible way to understand the disabilities and the needs of service users in community services with the multi-disciplinary care model.

Therefore, the purpose of this study was to adapt and validate a Hong Kong Chinese version of WHODAS 2.0 for persons with disabilities receiving community services in Hong Kong.

## Methods

### Subject recruitment

A convenient sampling technique was used. 177 clients with disabilities were recruited from four units subsidized by the Social Welfare Department SWD, Hong Kong Government, and held by a local non-governmental organization (NGO). The total service boundaries covered by the four units were around 48% of persons with disabilities in Hong Kong [18].

The following types of disabilities were included in the study: (1) physical disability; (2) hearing impairment; (3) visual impairment; (4) speech impairment; (5) mental illness; (6) intellectual disability (ID); (7) autism, attention deficit hyperactivity disorder (ADHD) and specific learning difficulties; and (8) chronic illness/visceral disability.

Inclusion criteria were: (1) be able to have basic communication and understandings the questions; (2) having no major hearing difficulties; (3) be willing to participate in the study; (4) be active users having at least one service in one month at the corresponding unit [19]. Exclusion criteria were: (1) with a diagnosis of moderate and severe ID; (2) severe hearing impairment. All subjects were informed about the purposes, the procedures and the confidentiality of the study. An information sheet of the study was distributed to them and all were required to sign a consent form for the written consent. A copy of the written consent form was given to each participant. For vulnerable subjects, their parents or caregivers were invited during the consent explanation and they acted as the witness of signing the written consent.

The subjects who agreed to participate in this study, were also invited for test-retest reliability using convenient sampling within 2 weeks.

For the recruitment procedures, potential participants were recruited from four rehabilitation units from the “Christian Family Service Centre (CFSC)”, which serves the community with the motto “Improving quality of life, uniting clients and community”.

The first unit was “Community Rehabilitation Day Centre (KRD)”, which mainly serves clients with physical disabilities and chronic illnesses living in Kowloon, Hong Kong. It provided exercise training on a day centre basis. Active users were 225, and 14 subjects were recruited in KRD.

The second unit was “Cheerful Place - District Support Centre - Kwun Tong East (DSC)” which provided day care service and day respite care on a centre basis for clients with ADHD, ID, speech impairment, and visual impairment living in Kowloon East, Hong Kong. Active users were 450, and 33 subjects were recruited in DSC.

The third unit was “Everjoy (RHCS)” which offers home care services for clients with severe disabilities on an outreach basis in their home. Rehabilitation services, personal care services, and nursing care were provided for those with physical disability, ID, ADHD, and chronic illness/visceral disability living in Kowloon East, Hong Kong. Active users were 550, and 88 subjects were recruited in RHCS.

The last unit was an outreach team for private residential care homes (POT) serving clients with mental illness, physical disability, ID, ADHD, hearing impairment, and visual impairment. Rehabilitation services and recreational activities were provided for these clients in private hostels in Kowloon and Hong Kong Island, Hong Kong. Active users were 280, and 42 subjects were recruited in POT.

The study period was from January 2023 to December 2024. The official permission to use WHODAS 2.0 was granted by WHO in April 2025. The study was approved by the Research and Ethics Committee of the Caritas Institute of Higher Education, Hong Kong (HRE220182).

### Translation process

The initial phase of the study included translation and adaptation of the WHODAS 2.0 from its original version in Hong Kong Chinese, the commonest form/dialect of spoken Chinese used by the Hong Kong population.

The standard “forward-backward” procedure was adopted to translate the English WHODAS 2.0 to Hong Kong Chinese to ensure both the accuracy of meaning and cultural acceptability. The forward translation was performed in close collaboration with experienced healthcare professionals in the care of clients with disabilities. This version was blindly back-translated to English by a bilingual native English speaker with a healthcare background.

The two versions of the WHODAS 2.0 were then assessed and compared by a group of healthcare professionals experienced in the care of clients with disabilities and independent linguistic experts. These experts modified the Hong Kong Chinese version when there were differences in meanings between the two versions until a final working Hong Kong Chinese version was approved.

A pilot test of the WHODAS 2.0 TC-HK was completed on 10 clients with disabilities to make sure they could understand and complete all the items.

### Data collection

The second phase of the study involved data collection to examine the validity and reliability of the WHODAS 2.0 TC-HK in persons with disabilities undergoing Community Rehabilitation Services.

### Demographics and disability characteristics

Participants’ characteristics, including age, gender, marital status, education level, and employment status, while disability characteristics, including types of diagnosed disabilities and the presence of co-morbidity (more than one type of disability), were collected for further data analysis.

During the rehabilitation training, participants were asked to complete the WHODAS 2.0 TC-HK, Hong Kong Chinese version of 36-Item Short Form Health Survey (SF-36 TC-HK), and Hong Kong Chinese version of the World Health Organization Quality of Life Abbreviated (WHOQOL-BREF TC-HK).

### Hong Kong Chinese version of World Health Organization Disability Assessment Schedule (WHODAS 2.0 TC-HK)

WHODAS 2.0 TC-HK, a 36-item self-administered version, is an instrument to assess functioning in six life domains: Understanding and communicating (6 items), Getting around (5 items), Self-care (4 items), Getting along with people (5 items), Life activities and Participation in society (8 items). Participants were required to score their level of difficulty in the past 30 days using a 5-point scale (0= no difficulty to 4 = extreme difficulty or cannot do). The higher the score, the greater disability was.

### Hong Kong Chinese version of 36-Item Short Form Health Survey (SF-36 TC-HK)

SF-36 TC-HK is to measure generic health status and has been widely used in the past two decades in local clinical settings [20–21]. A 36-item self-report measure of health-related quality of life (HRQOL) was also collected. Eight subscales are measuring different domains of HRQOL: physical functioning (PF), role-physical (RP), bodily pain (BP), general health (GH), vitality (VT), social functioning (SF), role-emotional (RE), and mental health (MH). The higher the scores, the better the health and functioning.

### Hong Kong Chinese version of the World Health Organization Quality of Life Abbreviated (WHOQOL-BREF TC-HK)

The WHOQOL-BREF, an abbreviated version of the World Health Organization Quality of Life (WHOQOL)-100 instrument, was also collected. It was developed by WHO to be the generic quality of life assessment [22]. WHOQOL-BREF TC-HK was commonly used to measure quality of life in local studies and acted as the gold standard [23,24]. It consists of 26 questions covering four quality-of-life domains: physical (seven items), psychological (six items), social (three items), and environmental (eight items), with two more general questions about health and quality of life. It has a Likert response scale varying from 1 to 5, and scores for each domain, as well as a total score, can be calculated. These scores were represented along a linear scale from 0 to 100, where higher scores reflect better quality of life.

### Statistical analysis

Descriptive statistics were used to summarize demographic data. Each score in outcome measures was calculated based on the formula.

The IBM SPSS Statistics for Windows, version 27.0 (IBM Corporation, Armonk, NY) was utilized for data analysis. The level of statistical significance was set at p≤0.05.

To assess convergent validity, Spearman’s correlation coefficient (rs) was used to investigate the associations between WHODAS 2.0 TC-HK and SF-36 TC-HK. To assess concurrent validity, Spearman’s correlation coefficient (rs) was used to investigate the associations between WHODAS 2.0 TC-HK and WHOQOL-BREF TC-HK. The interpretation rs value was lower than 0.25 as small, 0.25-0.50 as moderate, 0.50-0.75 as good; higher than 0.75 as excellent [25]. To assess reliability, the internal consistency of the WHODAS 2.0 TC-HK was estimated by Cronbach’s alpha coefficient; an alpha score of 0.8 or above was considered good [26]. Test-retest reliability was assessed by Intraclass correlation coefficient (ICC),0.5-0.75 as moderate reliability, 0.75-0.90 as good reliability, and higher than 0.9 indicating excellent reliability [27]. Furthermore, the construct validity of the WHODAS 2.0 TC-HK was tested by confirmatory factor analysis (CFA) to check whether the original hypothesised structure of the WHODAS 2.0 was fitted. To define a satisfactory fit of the model, the cut-off of root means square error of approximation (RMSEA), Goodness of Fit Index (GFI), and Standardized Root Mean Square Residual (SRMR) should be lower than 0.08, higher than 0.90, and lower than 0.08 respectively [28–30]. For the item factor loadings in CFA, it was analysed as ≥0.71 (excellent), 0.63-0.70 (very good), 0.55-0.62 (good), 0.45-0.54 (fair), 0.32-0.44 (poor), and<0.32 (unacceptable and deleted from factor) [31].

## Results

Data from 177 participants were collected and analysed, with subgroups categorized as KRD (n=14), DSC (n=33), RHCS (n=88), and POT (n=42). Thirty-six participants completed the WHODAS 2.0 TC-HK twice in two different sessions to evaluate the test-retest reliability. Table 1 showed the detailed distribution of age, gender, education, work status, and types of disabilities. The mean age of participants was 55 years (SD ±15.1). The RHCS subgroup had the oldest participants (60.9 ±14.5 years), while the DSC group was youngest (43.7 ±13.6 years). A slightly higher proportion was female in the samples (53.1%). About 32.2% (n=57) of participants were living independently in the community. All POT participants were hospitalized, contrasting with the DSC group, where 57.6% lived independently. The mean education level was 10.6 years (SD ±5.2).

**Table 1:** Participants’ demographics and types of disabilities.

|  | Total Sample<br>(n=177) | KRD (n=14) | DSC (n=33) | RHCS (n=88) | POT (n=42) |
| --- | --- | --- | --- | --- | --- |
| Age, Mean (SD) | 55.2±(15.1) | 52.9±(14.5) | 43.7±(13.6) | 60.9±(14.5) | 53.1±(11.7) |
| Female | 94 (53.1%) | 8 (57.1%) | 15 (45.5%) | 49 (55.7%) | 22 (52.4%) |
| Male | 83 (46.9%) | 6 (42.9%) | 18 (54.5%) | 39 (44.3%) | 20 (47.6%) |
| **Living condition** |  |  |  |  |  |
| Independent in<br>community | 57 (32.2%) | 7 (50%) | 19 (57.6) | 31(35.2%) | 0 |
| Assisted living | 77 (43.5%) | 7 (50%) | 13 (39.4%) | 57(64.8%) | 0 |
| Hospitalized | 43 (24.3%) | 0 | 1 (3.0%) | 0 | 42 (100%) |
| Education years,<br>Mean(SD) | 10.6±(5.2) | 12.4±(5.1) | 12.1±(5.0) | 9.6±(5.5) | 11.1±(4.5) |
| **Work status** |  |  |  |  |  |
| Paid Work | 20 (11.3%) | 0 | 10 (30.3%) | 5 (5.7%) | 5 (11.9%) |
| Unemployed other<br>reason | 6 (3.4%) | 0 | 4 (12.1%) | 0 | 2 (4.8%) |
| Unemployed health<br>reason | 56 (31.6%) | 8(57.1%) | 8 (24.2%) | 28 (31.8%) | 12 (28.6%) |
| Student | 2 (1.1%) | 0 | 1 (3.0%) | 1 (1.1%) | 0 |
| Keeping house | 10 (5.6%) | 1(7.1%) | 1 (3.0%) | 7 (8.0%) | 1 (2.4%) |
| Retired | 58 (32.8%) | 4(28.6%) | 2 (6.1%) | 39 (44.3%) | 13 (31.0%) |
| Non-paid work | 2 (1.1%) | 0 | 0 | 0 | 2 (4.8%) |
| Self employed | 2 (1.1%) | 1(7.1%) | 0 | 0 | 1 (2.4%) |
| Other | 21 (11.9%) | 0 | 7 (21.2%) | 8 (9.1%) | 6 (14.3%) |
| **Disability** |  |  |  |  |  |
| Physical disability | 49 (27.7%) | 10 (71.4%) | 13 (39.4%) | 25 (28.4%) | 1 (2.4%) |
| Hearing impairment | 10 (5.6%) | 0 | 2 (6.1%) | 7 (8.0%) | 1 (2.4%) |
| Visual impairment | 16 (9.0%) | 0 | 0 | 12 (13.6%) | 4 (9.5%) |
| Speech impairment | 13 (7.3%) | 4 (28.6%) | 1 (3.0%) | 6 (6.8%) | 2 (4.8%) |
| Mental illness | 31 (17.5%) | 0 | 1 (3.0%) | 2 (2.3%) | 28 (66.7%) |
| Intellectual disability | 14 (7.9%) | 0 | 8 (24.2%) | 2 (2.3%) | 4 (9.5%) |
| ADHD | 10 (5.6%) | 0 | 5 (15.2%) | 4 (4.5%) | 1 (2.4%) |
| Chronic illness/visceral disability | 34 (19.2%) | 0 | 3 (9.1%) | 30 (34.1%) | 1 (2.4%) |

Regarding work status, Unemployment due to health reasons was prevalent in KRD (57.1%) and RHCS (31.8%), whereas 30.3% of the DSC group maintained paid employment.

Physical disabilities were the most common, affecting 27.6% (n=49) of participants, with the highest prevalence in the KRD subgroup (71.4%). Mental illness was reported by 17.5% (n=31) of participants, with the POT subgroup showing the highest prevalence (66.7%). Chronic illnesses or visceral disabilities affected 19.2% (n=34) of participants, primarily in the RHCS subgroup (34.1%).

The test–retest period was 1–16 days, and the mean day was 5.9. 24 clients completed the 32-item version (Do5(2): Life activities: work/study domain cannot be filled) while 12 clients completed the 36-item version. Participants’ demographics and types of disabilities were shown in S1 Table.

Summary scores and reliability of the WHODAS 2.0 TC-HK, SF-36 TC-HK, and WHOQOL-BREF TC-HK were shown in Table 2. WHODAS 2.0 TC-HK domains’ medians ranged from 25 (Do1: Cognition) to 50 (Do2: Mobility, Do5(1): Life Activities – Household), with interquartile ranges (IQRs) varying between 30–60, indicating moderate to high disability levels. SF-36 TC-HK scores showed notable variability, ranging from 25 to 66.67 for role limitations due to physical health (IQR: 75) and role limitations due to emotional problems (IQR: 100), implying the severe effect from the physical aspect and the mild effect from the emotional aspect on role activities. WHOQOL-BREF TC-HK medians were 50 (Physical, Psychological) and 58.33–59.38 (Social, Environment), with narrow IQRs (16.67-23.21), suggesting less variability in quality-of-life perceptions.

**Table 2:** Summary scores and reliability of the WHODAS 2.0 TC-HK, Summary scores of SF-36 TC-HK, and WHOQOL-BREF TC-HK.

| Domain | N | Median | IQR | Cronbach's Alpha | Test-retest ICC[a] |
| --- | --- | --- | --- | --- | --- |
| **WHODAS 2.0 TC-HK (the higher the scores, the more disability)** |  |  |  |  |  |
| Do1: Cognition | 177 | 25 | 30 | 0.883 | 0.591 |
| Do2: Mobility | 177 | 50 | 59.35 | 0.886 | 0.872 |
| Do3: Self-care | 177 | 30 | 40 | 0.873 | 0.807 |
| Do4: Getting along | 177 | 33.3 | 33.3 | 0.883 | 0.669 |
| Do5(1): Life activities: household | 177 | 50 | 60 | 0.857 | 0.765 |
| Do5(2): Life activities: work/study | 48 | 28.6 | 35.7 | 0.875 | 0.798[b] |
| Do6: Participation | 177 | 37.5 | 33.4 | 0.869 | 0.815 |
| **SF-36 TC-HK (the lower the scores, the more disability)** |  |  |  |  |  |
| Physical functioning | 177 | 45 | 45 |  |  |
| Role limitations due to physical health | 177 | 25 | 75 |  |  |
| Role limitations due to emotional problems | 177 | 66.67 | 100 |  |  |
| Energy/fatigue | 177 | 50 | 25 |  |  |
| Emotional well-being | 177 | 64 | 26 |  |  |
| Social functioning | 177 | 50 | 37.5 |  |  |
| Pain | 177 | 57.5 | 38.75 |  |  |
| General health | 177 | 45 | 30 |  |  |
| Health change | 177 | 50 | 50 |  |  |
| **WHOQOL-BREF TC-HK (the lower the scores, the more disability)** |  |  |  |  |  |
| Physical | 177 | 50 | 23.21 |  |  |
| Psychological | 177 | 50 | 25 |  |  |
| Social Relationships | 177 | 58.33 | 16.67 |  |  |
| Environment | 177 | 59.38 | 18.75 |  |  |

[a]For test-retest, there were 36 patients analyzed in the study
[b]For Life activities: work/study, there were 12 patients analyzed for ICC

All WHODAS 2.0 TC-HK domains demonstrated high internal consistency, with Cronbach’s alpha scores ranging from 0.86 to 0.89, indicating strong reliability from the cut-off of satisfactory score as above 0.8. The highest consistency was observed in the mobility (α = 0.89) and cognition domain (α = 0.88). The lowest, though still excellent, was for life activities: household domain (α = 0.86).

WHODAS2.0 TC-HK has shown moderate to good test-retest reliability (ICC=0.59-0.87). The highest reliability was found in mobility (ICC = 0.87) and the participation domain (ICC = 0.82). The lowest was the cognition domain (ICC = 0.59), suggesting some response variability over time. For life activities: work/study domain, which was analysed in 12 patients, ICC was 0.80, indicating good reliability despite the smaller sample.

From Table 3, WHODAS2.0 TC-HK domains and SF-36 TC-HK sub scores were negatively and moderately correlated with statistical significance (rs = -0.25 to -0.81, p < 0.05) in most of the items. For the correlations between WHODAS2.0 TC-HK domains and WHOQOL-BREF sub scores, significantly negative and moderate correlations were found as rs = -0.29 to -0.62 in majority of items (Table 4).

**Table 3.** Spearman’s correlation coefficients between WHODAS2.0 TC-HK and SF-36 TC-HK.

| **WHODAS2.0**<br>**TC-HK**<br>**domain/SF-36**<br>**TC-HK sub-**<br>**score** | **Cognition** | **Mobility** | **Self-**<br>**care** | **Getting**<br>**along** | **Life activities**<br>**(domestic**<br>**responsibilities)** | **Life activities**<br>**(work &**<br>**school)** | **Participation** |
| --- | --- | --- | --- | --- | --- | --- | --- |
| **Physical functioning** | -0.003 | -0.805** | -0.599** | -0.188* | -0.522** | -0.590** | -0.370** |
| **Role limitations due to physical health** | -0.111 | -0.405** | -0.262** | -0.115 | -0.345** | -0.310* | -0.400** |
| **Role limitations due to emotional problems** | -0.183* | -0.110 | -0.042 | 0.004 | -0.176* | -0.181 | -0.346** |
| **Energy/fatigue** | -0.299** | -0.295** | -0.191* | -0.203** | -0.322** | -0.173 | -0.480** |
| **Emotional well being** | -0.363** | -0.069 | -0.080 | -0.147 | -0.140 | -0.140 | -0.382** |
| **Social functioning** | -0.279** | -0.294** | -0.199** | -0.186* | -0.239** | -0.241 | -0.530** |
| **Pain** | -0.131 | -0.268** | -0.095 | 0.034 | -0.134 | -0.233 | -0.250** |
| **General health** | -0.231** | -0.283** | -0.265** | -0.204** | -0.166* | -0.287* | -0.390** |
| **Health change** | -0.157* | -0.199** | -0.147 | -0.022 | -0.149* | -0.411** | -0.200** |

\*\* . Correlation is significant at the 0.01 level (2-tailed).
\*. Correlation is significant at the 0.05 level (2-tailed).

**Table 4.** Spearman’s correlation coefficients between WHODAS2.0 TC-HK and WHOQOL-BREF TC-HK.

| **WHODAS2.0**<br>**TC-HK**<br>**domain/**<br>**WHOQOL-**<br>**BREF TC-**<br>**HK** | **Cognition** | **Mobility** | **Self-care** | **Getting**<br>**along** | **Life activities**<br>**(domestic**<br>**responsibilities)** | **Life**<br>**activities**<br>**(work &**<br>**school)** | **Participation** |
| --- | --- | --- | --- | --- | --- | --- | --- |
| **Physical**<br>**health** | -0.349** | -0.549** | -0.491** | -0.386** | -0.586** | -0.469** | -0.618** |
| **Psychological**<br>**health** | -0.399** | -0.139 | -0.168* | -0.297** | -0.329** | -0.318* | -0.432** |
| **Social**<br>**relationships** | -0.313** | -0.072 | -0.154* | -0.389** | -0.291** | -0.263 | -0.371** |
| **Environment** | -0.316** | -0.156* | -0.184* | -0.301** | -0.225** | -0.176 | -0.414** |

\*\* . Correlation is significant at the 0.01 level (2-tailed).
\*. Correlation is significant at the 0.05 level (2-tailed).

WHODAS2.0 TC-HK 32-item version, excluding items from Do5(2): Life activities: work/study, was tested in CFA for construct validity. A second-order 6-factor model with the standardized parameter estimates and fit indices was summarized (Fig. 1).

**Fig 1.**
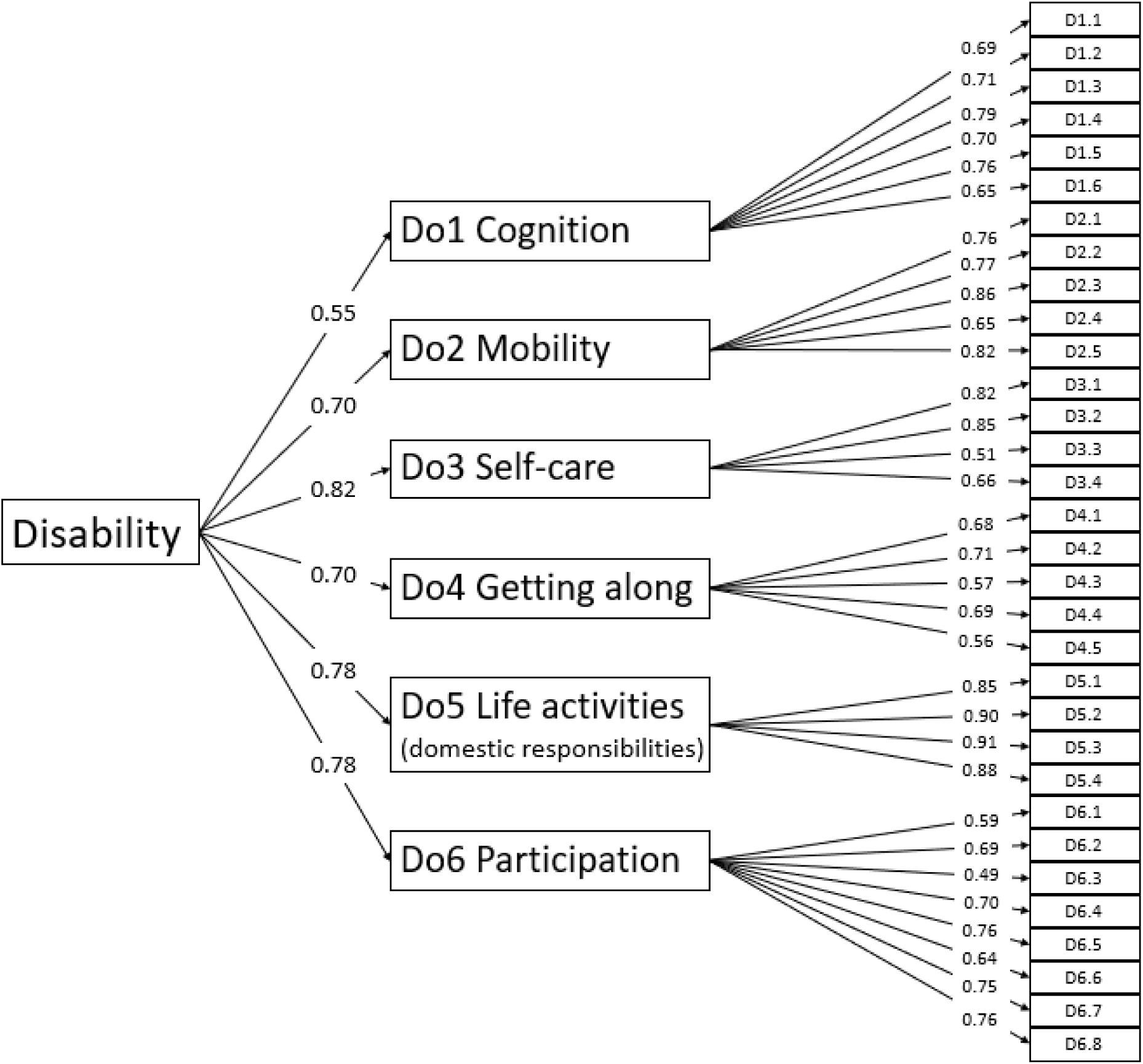
**Confirmatory factor analysis of WHODAS 2.0 TC-HK 32 items, model fit indicators(n=177): RMSEA = 0.086, GFI = 0.733, SRMR = 0.105**

## Discussion

Based on our best knowledge, the present study was the first to develop and validate the Hong Kong Chinese version of WHODAS 2.0. The properties measured in WHODAS 2.0 TC-HK were tested to be a valid and reliable tool in assessing disability for persons with disabilities in the Hong Kong community settings.

Satisfactory internal consistency was found as Cronbach’s alpha was all above 0.8. This was consistent with other studies with similar subjects [8,11,14]. Five domains showed good test-retest reliability (0.77-0.87). The lowest ICC was found in Do 1 Cognition (0.59) and Do 4 Getting along (0.67). In other studies, in Europe, Taiwan, and Norway, the ICC ranged from 0.20-0.74, 0.83-0.89, and 0.63-0.87, respectively [8,9,32]. As ICC was influenced by variability, it was found that Taiwan’s study was using face-to-face interviews in data collection, while the current study, Europe’s study, and Norway’s study were using self-administered questionnaires in data collection [8,9,32]. A similar study in Singapore showed that the ICC in the self-administered version would have a lower value [33]. Moreover, one of the drawbacks in the current study was that the sample size for reproducibility could not reach the recommended minimum(n=50) [34]. However, regarding the heterogeneity of the current study population, reaching a cut-off of 0.7 for all domains in Cronbach’s alpha and five out of seven domains in ICC indicated acceptable reproducibility for group comparisons in WHODAS2.0 TC-HK [8,27].

This study showed that WHODAS2.0 TC-HK had moderate to good convergent validity with SF36-TC-HK. The majority of domains of the 36-item WHODAS2.0 TC-HK were negatively correlated with domains of the SF-36 TC-HK. The higher the scores (the higher disability) at WHODAS2.0 TC-HK indicated the lower the scores (the lower quality of life) at SF36 TC-HK. The sub-scores of SF36-TC HK almost all have moderate correlation (rs=-0.25 to -0.53) with participation of WHODAS2.0 TC-HK. It showed the same with other similar studies [8–10]. This was coherent with the concept of ICF as participation being essential in accessing disability [1–2,4].

Furthermore, WHODAS2.0 TC-HK had moderate to good concurrent validity with WHOQOL-BREF TC-HK. WHODAS2.0 TC-HK was negatively and moderately correlated with WHOQOL-BREF TC-HK, as 19 out of 28 correlations rs =-0.29 to -0.57 and p<0.05. Compared with other studies, a similar result was obtained in Taiwan [32]. It demonstrated that WHODAS2.0 TC-HK was valid and valuable practically. However, it could not express all the meanings in WHOQOL-BREF TC-HK. Indeed, the result was quite consistent with the original study during the development of WHODAS 2.0 in 2010 [6].

For the CFA model, WHODAS2.0 TC-HK 32-item version was used as there was 48 participants (27.1%) filled in domain Do5(2): Life activities: work/study. Similar study conducted in Indonesia had 25 out of 183 participants (13.7%) completing domain Do5(2) [11]. It was coherent to the statistic done by the Hong Kong government showing that 79.9% persons with disabilities and chronic diseases were economically inactive [18]. The CFA model did not reach a satisfactory fit, which indicated some misfit. There were similar findings in the Indonesian version and the Norwegian version [8,11]. From the result, all factor loadings were higher than 0.45 (fair), with the majority higher than 0.54. The lowest was item 6.3 (0.49). A similar finding was shown in Taiwan Chinese population [32]. Item 6.3 was related to dignity. Given the cultural context between Chinese and English populations, there were substantial differences in how to define dignity, as it may include teamwork, initiative, autonomy, and respect for elders [35]. Persons with disabilities experienced loneliness, low perceived social support, and social isolation as they experienced more barriers in the physical environment and social participation [36–37]. These may affect how they perceive dignity in the past 30 days.

There were some limitations in the present study. Sampling size was relatively small when comparing similar studies [8,13,38]. Based on the guideline proposed in 2016, sample sizes less than 100 cases should not be recommended for any type of Structural Equation Modeling [39]. While the sample-to-item ratio should not be less than 5:1 [40]. Therefore, in this study, N=177 should be considered as the acceptable level. More importantly, it was suggested that a large sample size with blind selection (N > 300) was not more meaningful than a small sample size with careful selection (N > 150) [41]. Secondly, participants recruited were from two geographical regions of Hong Kong and four units held by one NGO. The generalizability of our findings to others living in different regions and receiving services from different NGOs may need further studies. Another limitation was that the assessment tools were in Hong Kong Traditional Chinese. Therefore, for those who were not fluent in Cantonese, the validity and reliability may not be applicable. As 90.6% of the population in Hong Kong were using Cantonese [42], the current findings were useful and applicable to the service users in Hong Kong.

## Conclusions

WHODAS2.0 TC-HK is a reliable and valid tool in assessing persons with disabilities who receive community services in Hong Kong. Despite the limitations mentioned above, the results showed satisfactory reliability and moderate convergent and concurrent validity. As WHODAS2.0 TC-HK is being generic and easy to administer, it can be considered as the first-choice instrument in the community rehabilitation field for collecting and developing the ICF database in Hong Kong. Further studies are needed to evaluate the short- and long-term effectiveness and efficiency of WHODAS 2.0 TC-HK implemented in clinical settings of social welfare services.

## Data Availability

Identifiable personal details of all subjects were encoded during data collection. Only the permitted researchers were having the key for decoding. The completed questionnaires were stored confidentially at the S. K. Yee School of Health Sciences, Saint Francis University Hong Kong. The datasets analyzed in the current study are not publicly available because of privacy issue. Please send email to for educational request.

## Acknowledgements

Thank you for the WHO’s official permission of using WHODAS 2.0. Moreover, this was our honour to be supported by the recruited subjects to participate this research. They showed their eagerness to provide the information for us so as to recognize the newly implemented assessment tool. Also, we would like to thank the staff working in Christian Family Service Centre in subject recruitment and data collection.

## Supporting information

**S1Table. Participants’ demographics and types of disabilities for test-retest.** (DOCX)

